## Supplementary figures and tables for "Risk of COVID-19 death in adults who received booster COVID-19 vaccinations: national retrospective cohort study on 14.6 million people in England"

**Supplementary Figure 1** – Population diagram

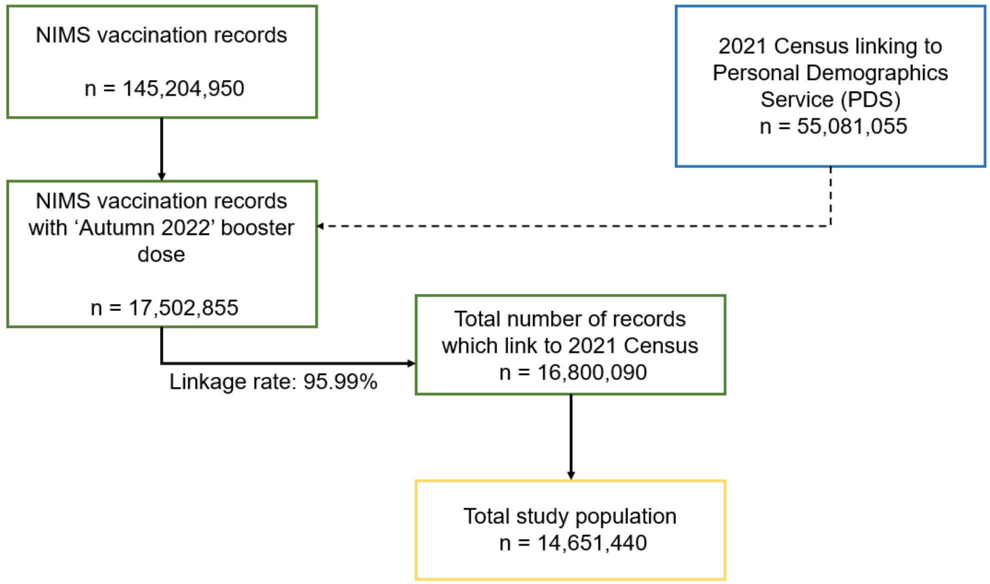

***Supplementary Table 1*** *– Population sample flow*

| Stage | Total population |
|---|---|
| Total 2021 Census linking to Personal Demographics Service (PDS) | 55,081,055 |
| Total number of NIMS records | 145,204,950 |
| Total number of NIMS records with autumn 2022 dose | 17,502,855 |
| Total number of NIMS records with autumn 2022 dose which link to 2021 Census | 16,800,090 |
| Total population who were usual residents at time of 2021 Census | 16,777,280 |
| Total population aged 50 to 100 years of age on vaccination date and alive on 1 September 2022 | 14,657,455 |
| Total people with positive time at risk (time between 14 days after vaccination and the earliest of end of study (11 April 2023) date or date of death) | 14,651,440 |

**Supplementary Table 2** – Z-statistics estimates for health predictors from bootstrapped estimates

| **Category** | **Z-statistic** |
|---|---|
| Asthma | 1.32 |
| Atrial Fibrillation | -0.32 |
| Cancer of Blood or Bone Marrow | 5.96 |
| Chronic Kidney Disease | 3.06 |
| Congenital Heart Problem | -0.93 |
| Chronic obstructive pulmonary disease (COPD) | 0.54 |
| Coronary Heart Disease | 1.07 |
| Cystic fibrosis * | 3.47 |
| Dementia | -7.15 |
| Diabetes: Type 1 | 0.03 |
| Diabetes: Type 2 | -1.02 |
| Epilepsy | -0.69 |
| Heart Failure | 0.79 |
| Immunosuppressed | 1.26 |
| Learning Disability or Down Syndrome | 1.15 |
| Prescribed leukotriene | 0.98 |
| Liver Cirrhosis | -1.86 |
| Lung or Oral Cancer | -0.04 |
| Motor neurone disease /Multiple sclerosis/Myasthenia/Huntington’s/Chorea | 0.23 |
| Parkinson’s Disease | 0.36 |
| Peripheral Vascular Disease | -1.58 |
| Prior Fracture *** | -1.16 |
| Pulmonary Hypertension or Fibrosis | 2.45 |
| Rheumatoid Arthritis or Systemic lupus erythematosus (SLE) | 2.77 |
| Schizophrenia | 0.13 |
| Severe mental illness | 0.21 |
| Stroke ** | -0.07 |
| Thrombosis or pulmonary embolus | 0.34 |

* - Cystic fibrosis, bronchiectasis or alveolitis, ** - Stroke or transient ischaemic attack (TIA), *** - Prior fracture of hip, wrist, spine, or humerus

***Supplementary Table 3*** – Health conditions associated with COVID-19 and non-COVID-19 death from model estimates not adjusted for all other health conditions

| **Category** | **HR for COVID-19 death (95% CI)** | **HR for non-COVID-19 death (95% CI)** |
|---|---|---|
| Asthma | 1.21 (1.1 - 1.33) | 1.07 (1.05 - 1.09) |
| Atrial Fibrillation | 1.6 (1.5 - 1.71) | 1.58 (1.56 - 1.6) |
| Cancer of Blood or Bone Marrow | 3.58 (3.14 - 4.07) | 1.9 (1.83 - 1.97) |
| Chronic Kidney Disease | 1.55 (1.46 - 1.65) | 1.38 (1.36 - 1.4) |
| Congenital Heart Problem | 1.37 (0.78 - 2.41) | 1.59 (1.43 - 1.78) |
| Chronic obstructive pulmonary disease (COPD) | 2.51 (2.35 - 2.69) | 2.3 (2.27 - 2.34) |
| Coronary Heart Disease | 1.43 (1.32 - 1.53) | 1.33 (1.31 - 1.35) |
| Cystic fibrosis * | 2.5 (2.17 - 2.88) | 1.65 (1.59 - 1.72) |
| Dementia | 2.83 (2.63 - 3.04) | 3.66 (3.61 - 3.71) |
| Diabetes: Type 1 | 1.66 (1.48 - 1.85) | 1.68 (1.65 - 1.72) |
| Diabetes: Type 2 | 1.46 (1.37 - 1.55) | 1.51 (1.49 - 1.53) |
| Epilepsy | 1.81 (1.41 - 2.33) | 2.05 (1.95 - 2.15) |
| Heart Failure | 2.34 (2.18 - 2.52) | 2.19 (2.15 - 2.23) |
| Immunosuppressed | 2.62 (2.34 - 2.93) | 1.88 (1.83 - 1.93) |
| Learning Disability or Down Syndrome | 5.49 (4 - 7.53) | 4.72 (4.42 - 5.04) |
| Prescribed leukotriene | 1.79 (1.69 - 1.89) | 1.61 (1.59 - 1.63) |
| Liver Cirrhosis | 3.52 (2.6 - 4.77) | 4.48 (4.23 - 4.73) |
| Lung or Oral Cancer | 3.82 (3.03 - 4.8) | 3.66 (3.48 - 3.84) |
| Motor neurone disease /Multiple sclerosis/Myasthenia/Huntington's/Chorea | 3.44 (2.14 - 5.54) | 3.15 (2.84 - 3.5) |
| Parkinson's Disease | 3.08 (2.64 - 3.59) | 3.18 (3.08 - 3.29) |
| Peripheral Vascular Disease | 1.85 (1.61 - 2.13) | 2 (1.94 - 2.06) |
| Prior Fracture *** | 1.37 (0.95 - 1.97) | 1.62 (1.51 - 1.74) |
| Pulmonary Hypertension or Fibrosis | 4.09 (3.46 - 4.83) | 2.9 (2.78 - 3.03) |
| Rheumatoid Arthritis or Systemic lupus erythematosus (SLE) | 2.26 (1.95 - 2.62) | 1.52 (1.47 - 1.58) |
| Schizophrenia | 2.5 (1.89 - 3.31) | 2.45 (2.31 - 2.6) |
| Severe mental illness | 1.67 (1.52 - 1.83) | 1.66 (1.63 - 1.69) |
| Stroke ** | 1.56 (1.43 - 1.7) | 1.6 (1.58 - 1.63) |
| Thrombosis or pulmonary embolus | 2.53 (0.82 - 7.85) | 1.97 (1.49 - 2.59) |

* - Cystic fibrosis, bronchiectasis or alveolitis, ** - Stroke or transient ischaemic attack (TIA), *** - Prior fracture of hip, wrist, spine, or humerus
